# COVID-19 reinfection: A Rapid Systematic Review of Case Reports and Case Series

**DOI:** 10.1101/2021.03.22.21254081

**Authors:** Jingzhou Wang, Christopher Kaperak, Toshiro Sato, Atsushi Sakuraba

## Abstract

The COVID-19 pandemic has infected millions of people worldwide and many countries have been suffering from a large number of deaths. Acknowledging the ability of SARS-CoV-2 to mutate into distinct strains as an RNA virus and investigating its potential to cause reinfection is important for future health policy guidelines. It was thought that individuals who recovered from COVID-19 generate a robust immune response and develop protective immunity, however, since the first case of documented reinfection of COVID-19 in August 2020, there have been a number of cases with reinfection. Many cases are lacking genomic data of the two infections and it remains unclear whether they were caused by different strains. In the present study, we undertook a rapid systematic review to identify cases infected with different genetic strains of SARS-CoV-2 confirmed by polymerase-chain reaction and viral genome sequencing. A total of 17 cases of genetically confirmed COVID-19 reinfection were found. One immunocompromised patient had mild symptoms with the first infection, but developed severe symptoms resulting in death with the second infection. Overall, 68.8% (11/16) had similar severity, 18.8% (3/16) had worse symptoms, and 12.5% (2/16) had milder symptoms with the second episode. Our case series shows that reinfection with different strains is possible and some cases may experience more severe infections with the second episode. The findings also suggest that COVID-19 may continue to circulate even after achieving herd immunity through natural infection or vaccination suggesting the need for longer term transmission mitigation efforts.

---

Coronavirus disease 2019 (COVID-19) has infected over 38 million individuals and claimed at least one million lives across the globe since it originated in Wuhan, China in late 2019.(1) As an RNA virus that is prone to mutations, SARS-CoV-2 has been reported to have heterogeneous genetic composition in different geographical locations.(2) Since August 2020, several cases of COVID-19 reinfections have been reported. The current study aims to summarize these cases to facilitate our understanding on the degree of protective immunity.

Electronic databases (PubMed, MedRxiv, and Social Science Research Network) were searched from Jan 1^st^, 2020 to October 12^th^, 2020 using terms “SARS-CoV-2,” “CoV2,” “COVID-19,” and “reinfection”. COVID-19 reinfection was defined as individuals infected with different genetic strains of SARS-CoV-2 confirmed by polymerase-chain reaction. Only studies with viral genome sequencing available for both infectious events were included in this report to distinguish true re-infection and prolonged viral shedding, as research has shown that a certain proportion of patients may continue to carry the virus despite resolution of symptoms and prior negative PCR tests.(3) For this reason, six peer-reviewed articles and two news articles from the stated date range describing either individual or small groups of additional possible COVID-19 reinfections (totaling 31 individuals) were excluded. The present study was a rapid systematic review of published case reports or case series, so approval of Institutional Review Board (IRB) was not necessary.

A total of 17 cases of genetically confirmed COVID-19 reinfection have been reported in the literature to date, which are summarized in **Table 1**. Reinfection has been reported in Asia, Europe, and North and South America. Ages of reinfected individuals ranged between 24 and 89 years old. Mean interval between the first and the second infection averaged 76 days (range 19-142). Only one reinfected patient was immunocompromised (1/17, 5.8%). This patient was a female in her 80s undergoing chemotherapy for a hematologic malignancy, who had mild symptoms with her first infection, but developed severe symptoms resulting in death with her second infection.(4) Among the remaining 16 patients, the proportion of patients having mild/asymptomatic infections were the same for the first and second episodes (93.8%). Overall, 68.8% (11/16) had similar severity, 18.8% (3/16) had worse symptoms, and 12.5% (2/16) had milder symptoms with the second episode.

**Table 1:** Summary of COVID-19 Reinfection Cases with Confirmed Genomic Differences.

| Patient | Date reported | Age /sex | Immuno-compromised | Interval (days) | Symptom severity<sup>†</sup> (1st episode) | Symptom severity (2nd episode) | Negative PCR test in between infections | Seroconversion after initial infection | Seroconversion after second infection | Viral clade (1<sup>st</sup> episode) | Viral clade (2<sup>nd</sup> episode) | Recovered | Country | References<sup>‡</sup> |
| --- | --- | --- | --- | --- | --- | --- | --- | --- | --- | --- | --- | --- | --- | --- |
| 1 | May-20 | 42M | No | 51 | Mild | Mild | Not performed | Not performed | Not performed | B.1.26 | B.1.26 but with several mutations | Yes | U.S. | Larson *et al*, 2020<sup>§</sup> |
| 2 | Jun-20 | 25M | No | 48 | Mild | Moderate<sup>†</sup> | Yes | Not performed | Yes | 20C | 20C with 11 SNP mutations | Yes | U.S. | Tillett *et al*, 2020 |
| 3 | Jun-20 | 51F | No | 120 | Mild | Mild | Not performed | Not performed | Not performed | B.1.1 | A | Yes | Belgium | Van Elslande *et al*, 2020 |
| 4 | Jul-20 | 60M | No | 140 | Moderate | Mild | Yes | Not performed | Yes | 19B | 20A | Yes | U.S. | Goldman *et al*, 2020 |
| 5 | Jul-20 | 33M | No | 142 | Mild | Asymptomatic | Yes | Yes | Yes | D614 (V/19A;B.2) | G614 (G20A; B.1) | Yes | China | To *et al*, 2020 |
| 6 | Jul-20 | 46M | No | 68 | Mild | Mild | Yes | Not performed | Yes | B.1.p9 | A.1.1 | Yes | Ecuador | Prado *et al*, 2020 |
| 7 | Jul-20 | 89F | Yes | 59 | Mild | Severe<sup>†</sup> | Not performed | Not performed | No | N/A | 10 SNP mutations | No | The Netherlands | Mulder *et al*, 2020 |
| 8 | Jul-20 | 27M | No | 66 | Mild | Mild | Yes | Not performed | No | B.1 | B; 8 SNP mutations | Yes | India | Shastri *et al*, 2020 |
| 9 | Jul-20 | 31M | No | 65 | Asymptomatic | Mild<sup>†</sup> | Yes | Not performed | No | B.1.1 | B.1.1; 9 SNP mutations | Yes | India | Shastri *et al*, 2020 |
| 10 | Jul-20 | 27M | No | 19 | Asymptomatic | Mild<sup>†</sup> | Yes | Not performed | Not performed | B.1.1 | B.1.1, 9 SNP mutation | Yes | India | Shastri *et al*, 2020 |
| 11 | Jul-20 | 24F | No | 55 | Mild | Mild | Not performed | Not performed | No | B.1.1 | B.1.1, 12 SNP mutations | Yes | India | Shastri *et al*, 2020 |
| 12 | Aug-20 | 47F* | N/A | 88 | Mild | Mild | Not performed | Not performed | No | D614 | G614 | Yes | Qatar | Abu-Radda *et al*, 2020 |
| 13 | Aug-20 | 27M* | N/A | 46 | Mild | Mild | Not performed | Not performed | Not performed | D614 | G614 | Yes | Qatar | Abu-Radda *et al*, 2020 |
| 14 | Aug-20 | 42M* | N/A | 71 | Mild | Mild | Not performed | Not performed | Not performed | D614 | G614 | Yes | Qatar | Abu-Radda *et al*, 2020 |
| 15 | Aug-20 | 27M* | N/A | 55 | Mild | Mild | Not performed | Not performed | Not performed | D614 | G614 | Yes | Qatar | Abu-Radda *et al*, 2020 |
| 16 | Aug-20 | 25M | No | 106 | Asymptomatic | Asymptomatic | Yes | Not performed | Not performed | N/A | 9 SNP mutations | Yes | India | Gupta *et al*, 2020 |
| 17 | Sep-20 | 28F | No | 107 | Asymptomatic | Asymptomatic | Yes | Not performed | Not performed | N/A | 10 SNP mutations | Yes | India | Gupta *et al*, 2020 |

Individuals who recovered from COVID-19 were generally thought to generate a robust immune response to clear the virus. However, it remains to be determined whether the initial infection confers a protective immunity to subsequent infection(s). Recent research has suggested that positive COVID-19 antibody from initial infection may provide protection against reinfection in a majority of study participants, but reinfection is still possible in certain individuals. (5) Reinfection with other human coronaviruses is common, despite the presence of antibodies.(6) The current case series indicate that COVID-19 reinfection is possible and the second infection may result in worse symptoms in nearly 20% of patients and serious complications in those who are elderly and immunocompromised. Our data also suggests that reinfection is not specific to any particular strain and multiple strains with different genetic sequence have been shown to cause reinfection. Due to the emergence of the recently described spike deletion variants from UK and South Africa, it is of interest whether second infections can occur in people who have had COVID-19 during the “first wave” before these variants were prevalent.

Given the potential reporting bias and the current report only including studies with genomic data, there are likely many more reinfection cases than have been currently described. However, the true prevalence of COVID-19 reinfection may be difficult to estimate, considering that complete genomic data is not available in most COVID-19 infections and many patients with milder symptoms were not tested in the early phase of this pandemic. Additionally, people with asymptomatic reinfections are less likely to be identified, so identifying true prevalence of COVID-19 reinfection is difficult without population-based studies, which is a possible area for future research. Studies included in our analysis reported certain key nucleotide difference between the sequenced viruses, but more recently new variants have also been detected in areas of UK where cases are rising.(7) Some primary literature cited in our study did not contain data on seroconversion, so it is difficult to comment on the connection between immunity and presence of antibody, but one patient developed re-infection despite prior positive antibody test (patient 5). Considering that the two strains belong to the same clade in some reported cases, the possibility of accelerated mutation of the original strain or simultaneous infection with more than one strain in addition to waning immunity should be also considered. It is also difficult to differentiate between COVID-19 reinfection, relapse and PCR re-positivity in some cases and Yahav et al. proposed reinfection as >90 days apart(8), but we restricted our inclusion criteria to only patients with confirmed infection with different genetic strains. Two meta-analyses undertaken early in the pandemic reported that reinfection or re-positivity were rare, but lacked cases with genomic data.(9, 10)

Our case series indicate that previous COVID-19 exposure does not confer total immunity and that a second infection is possible. Therefore, individuals, regardless of history of prior infection, should continue to participate in mitigating the spread of infection by practicing social distancing and mask wearing. The findings also suggest that COVID-19 may continue to circulate in humans(11), even after achieving herd immunity through natural infection or vaccination.

## Data Availability

Data can be requested to the corresponding author.

